# Understanding good communication in ambulance pre-alerts to Emergency Department. Findings from a qualitative study of UK emergency services

**DOI:** 10.1101/2024.09.25.24314364

**Authors:** Fiona C. Sampson., Rachel O’Hara., Jaqui Long., Joanne Coster.

## Abstract

**Objectives:** Pre-hospital notifications (pre-alerts) enable Emergency Department (ED) staff to prepare for the arrival of patients requiring a time-critical response. Effective communication of the pre-alert is key to enabling the ED to prepare appropriately but evidence on communication practices is lacking. We undertook qualitative research to understand how pre-alert communication may be improved to optimise the ED response for pre-alerted patients.

**Design, setting and participants:** Data collection took place within three UK Ambulance Services and six EDs between August 2022-April 2023. We undertook semi-structured interviews with 34 ambulance and 40 ED staff and 156 hours non-participation observation of pre-alert practice (143 pre-alerts). Verbatim interview transcripts and observation notes were imported into NVivo™ and analysed using a thematic approach.

**Results:** We identified significant variation in how pre-alerts were communicated that influenced how effectively information was transferred. Ambulance and ED staff demonstrated a shared recognition that pre-alerts need to be communicated concisely, but both received minimal training in how to give and receive pre-alerts. Efficient pre-alerting was influenced by clinician experience and seniority. ED and ambulance clinicians following different information sharing formats (e.g. ATMIST, SBAR) sometimes led to interruptions, information loss and tensions, particularly when an early ‘headline’ clinical concern had not been shared. Ambulance clinicians sometimes questioned the appropriateness of their pre-alert when ED clinicians did not explain the rationale for not giving the expected response (i.e. being accepted into a high-priority area of ED). Additional sources of frustration included technological problems and poor communication of ETA and caller/responder identities.

**Conclusions:** Use of shared format including a headline ‘cause for concern’ may improve the clarity, usefulness and civility of pre-alerts, particularly when the clinician concern is not obvious from observations. Basic training on how to undertake pre-alerts for both ED and ambulance clinicians may improve understanding of the importance of pre-alert communication.

**Strengths and limitations of the study:**

- This study triangulated findings from Emergency Department staff and Ambulance Clinicians from across three Ambulance Service regions in England to provide insights into the causes of incivility relating to pre-alert communications.
- Semi-structured interviews and non-participant observation provide rich data regarding the experiences and practice of undertaking pre-alerts.
- Fieldwork took place within larger Emergency Departments (major trauma centres and trauma units) rather than minor units where pre-alerts occur less frequently and may be managed differently.
- The setting included only ambulance services where ambulance clinicians principally call directly to the ED, limiting transferability of findings for ambulance services who call via a control centre.
- Non-participant observation was undertaken in Emergency Departments but not within Ambulance Services due to the small number of pre-alerts occurring per shift.

## Introduction

Ambulance clinicians use pre-alert calls to Emergency Departments (EDs) to inform staff of the arrival of a critically ill or rapidly deteriorating patient who they believe will require time-critical treatment or senior clinical review upon arrival. This advance notification can facilitate earlier initiation of time-critical treatment, improved processes and better clinical outcomes by enabling EDs to prepare and have appropriate staff, medication and other resources available. (1-4) Pre-alerts are increasingly a key component of patient pathways for a range of conditions such as stroke, STEMI and major trauma (5-8).

However, pre-alerts are not risk-free, and inappropriate pre-alerting or action in response to pre-alerts may have an impact on other critically ill patients within the ED. This may be exacerbated in the current climate of high demand, ED crowding and long waiting times for ambulance handovers, with increasing pressures for both ED and ambulance staff.(6, 9) Concise communication of pre-alerts is essential to enable ED staff to prepare adequately and to minimise distraction of ambulance and ED clinicians from delivering patient care where it is most needed. Communication failures during handover of patient information are a recognised patient safety risk and identified as a top five World Health Organisation improvement priority.(10, 11) There is evidence of variation in the quality of information provided within pre-alerts, with high proportions of pre-alerts providing inaccurate, unnecessary or inadequate information to enable the EDs to appropriately mobilise resources. (12-14)

Despite the increasing body of evidence supporting the use of pre-alerts for patients requiring time-critical treatment, there is a lack of research exploring how pre-alerts should be communicated to maximise effectiveness. (15) Guidance explaining how pre-alerts should be undertaken is variable and Budd et al. reported that 40% of ambulance staff stated that they didn’t use standard content when alerting hospitals of incoming trauma. (13, 16)Similarly, a recent survey of UK ambulance clinicians identified significant differences in the use of mnemonics or structured formats during pre-alert. (17) As part of a wider mixed methods study exploring the use of pre-hospital pre-alerts, we aimed to understand how pre-alert information is communicated and how communication can be improved to optimise the ED response for pre-alerted patients.

## Methods

The study methods are reported according to the Standard for Reporting Qualitative Research (SRQR) criteria. Ethical approval was obtained from Newcastle & North Tyneside 2 Research Ethics Committee (Ref: 21/NE/0132).

### Patient and Public Involvement

Study design PPI input was provided via existing PPI contacts. A wider study PPI group including patients and carers with lived experience of pre-alerts was then established and met with the research team regularly throughout the study. The PPI group provided feedback on the data collection approach and documents, as well as the emerging findings. Their experience of pre-alerts helped to inform the ED observations. The findings were presented at an online PPI workshop to elicit views on the most important issues. Participant comments contributed to a wider stakeholder workshop on how the findings can be used to improve practice.

### Qualitative approach and research paradigm

We used a qualitative design based on a critical realist approach, in which we assume there is a shared and complex reality whose understanding can be described (albeit imperfectly) and is mediated by cultural contexts. The choice of qualitative methods was pragmatic and involved selecting the methods best suited to addressing the research problem - understanding how pre-alert information is communicated.(18) (19)

### Context

Fieldwork took place within three of the ten ambulance services (population approximately 15.5 million) in England who were selected to encompass a diversity of deprivation levels, rural/urban mix and diverse ethnic populations as well as high rates of electronic patient record (ePRF) completion by ambulance clinicians.

In the UK, trauma is managed via a network of major trauma centres (MTC), trauma units (TU) and other EDs. (20) The six ED study sites were selected by identifying one MTC and one TU per ambulance service who received high numbers of pre-alerts in order to ensure that sufficient pre-alert activity could be observed during the researcher visits.

### Sampling strategy

Ambulance clinicians were initially selected purposively, (21)sampling for length of experience, sex, role, ethnicity and whether they were high or low pre-alerters according to routine data. We recruited selected clinicians from direct invite before expanding recruitment via open invitations facilitated by research leads at each ambulance service. We also interviewed clinicians who we identified during observations at individual EDs (see below).

Interviews with ED staff and non-participant observations were undertaken at the six identified study sites. We recruited ED staff via direct invitation during observation and via local research leads who invited staff within particular roles (e.g. clinical leads). We aimed to sample a range of different roles at each site, including senior and junior medical and nursing staff as well as other roles identified as important at individual sites during the fieldwork (e.g. Hospital Ambulance Liaison Officers - HALOs). All participants were offered a £25 shopping voucher and CPD certificate as a thank-you for participating.

### Data collection methods

Observations and interviews were undertaken principally by two researchers (JL & JC) with FS also undertaking initial site visits. Observations took place predominantly within sight of the pre-alert phone (usually in the resuscitation area) but researchers also observed throughout the ED and the ambulance arrival and waiting areas. Staff were made aware of the presence of researchers and were given the option to opt out of being observed. Further details of how observations took place are available in supplementary file (1). Data collection continued until the minimum proposed number of interviews were undertaken and thematic saturation occurred during analysis. Interviews were conducted remotely, either online or by phone.

### Data collection instruments, technologies and processing

Interview topic guides were developed in collaboration with the project management and PPI group. Topic guides were used flexibly (i.e. all topics were covered, but not in the same order). Observation guides were developed and refined after initial visits, along with a form to record details of individual pre-alert calls (not recording any patient data). Interviews were recorded using encrypted dictaphones and transcribed verbatim. Data was stored in a restricted access university secure filestore, accessible only by the research team at University of Sheffield. All participants were allocated a unique code, which was used within data excerpts. Transcripts have not been made openly available to ensure anonymity.

All fieldwork data (interview transcripts and observation notes) were imported into NVivo ™ for analysis.

### Researcher characteristics and reflexivity

Observation notes were written up in detail shortly after the observations took place, including reflexive notes from the researchers. The researchers involved in data collection were all female, white, health service researchers with between 8 and 22 years experience and backgrounds in social science/psychology. Two of the researchers (FS and ROH) had prior experience of undertaking non-participant observation in emergency services settings. The field researchers (JL and JC) had no prior experience and had fewer pre-conceptions about how emergency services worked than ROH and FS.

### Data analysis

Data was analysed using a thematic approach according to the principles of Braun & Clark. (22) Data familiarisation involved ROH, JL, JC and FS reading a subset of the interview transcripts to develop initial themes and a preliminary coding framework. Coding was undertaken initially by ROH (who had not undertaken any fieldwork) and JL (who had conducted the majority of data collection). Data was coded independently and discussed within the wider research team on a weekly basis in order to refine coding and analysis. Changes to coding frameworks were documented and labelled at each stage. Code summaries were developed and cross-cutting themes identified after discussion during team meetings.

### Techniques to enhance trustworthiness

During the interviews, researchers summarised their understanding of the respondents’ accounts to clarify their interpretation. (23) Researcher triangulation within both the data collection and analysis phase helped improve trustworthiness of analysis. Findings were presented to PPI groups and at a national online workshop incorporating both a sample of research participants and key stakeholders from ambulance service and ED national bodies, where participants provided feedback.

### Details of interview participants and observations

We undertook interviews with a total of 34 ambulance clinicians from across the three ambulance services and 40 ED clinicians from the six EDs. Characteristics of respondents are detailed in tables 1 and 2 below:

**Table 1:** Details and number of participants per ambulance service.

|  |  |  |
| --- | --- | --- |
| Service | Site 1 | 13 |
|  | Site 2 | 11 |
|  | Site 3 | 10 |
| Role | Paramedic | 25 |
|  | Advanced/specialist paramedic | 6 |
|  | Technician | 2 |
|  | Clinical Lead | 1 |
| Years in role | <1 year | 6 |
|  | 1-5 years | 15 |
|  | 5-10 years | 9 |
|  | >10 years | 4 |
| Gender | Female | 13 |
|  | Male | 21 |
| Ethnicity | White British | 31 |
|  | Other ethnicity | 3 |

**Table 2:**
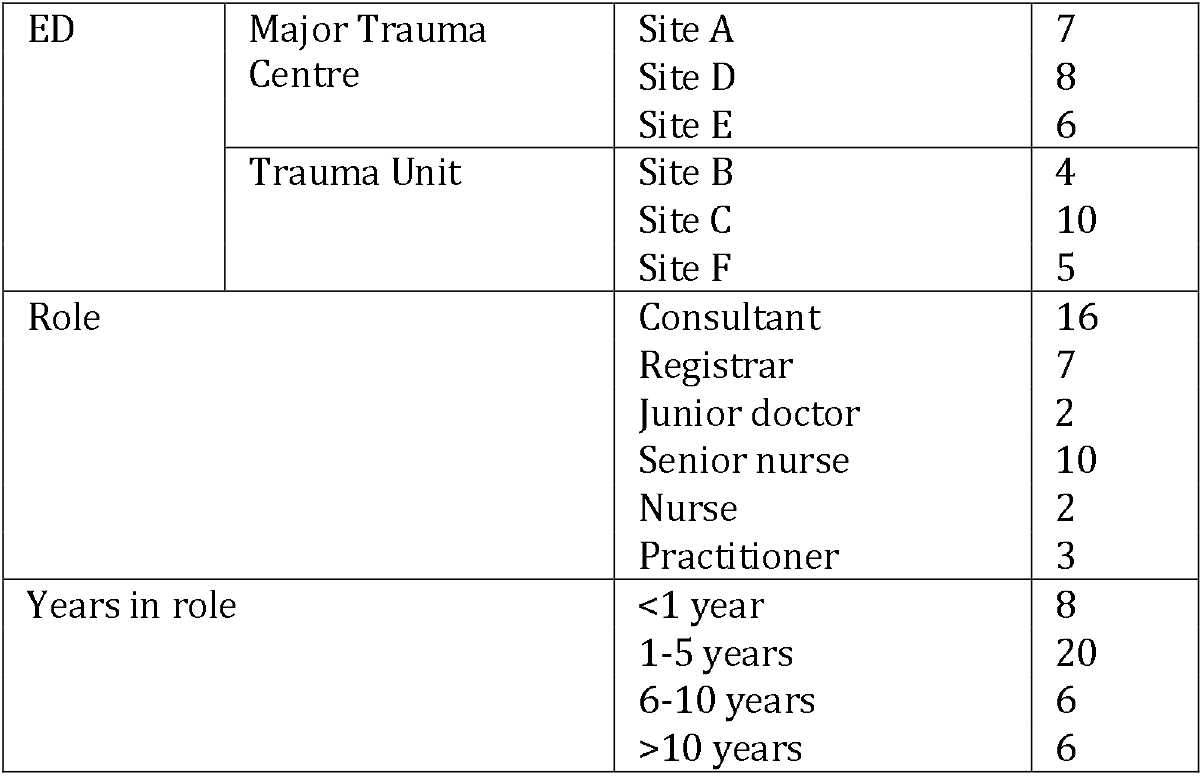
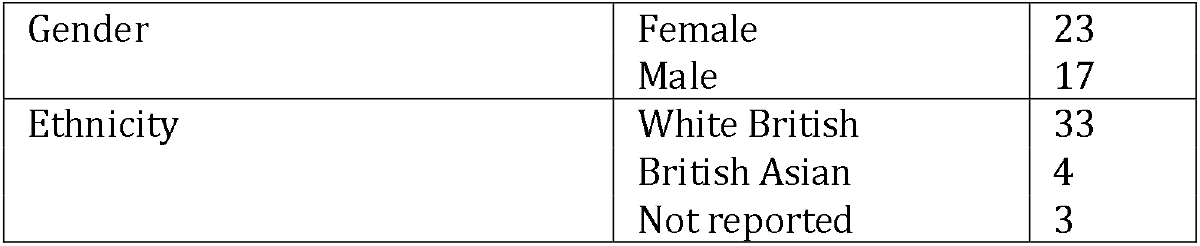
Details and number of staff participants per ED.

|  |  |  |  |
| --- | --- | --- | --- |
| ED | Major Trauma Centre | Site A | 7 |
|  |  | Site D | 8 |
|  |  | Site E | 6 |
|  | Trauma Unit | Site B | 4 |
|  |  | Site C | 10 |
|  |  | Site F | 5 |
| Role |  | Consultant | 16 |
|  |  | Registrar | 7 |
|  |  | Junior doctor | 2 |
|  |  | Senior nurse | 10 |
|  |  | Nurse | 2 |
|  |  | Practitioner | 3 |
| Years in role |  | <1 year | 8 |
|  |  | 1-5 years | 20 |
|  |  | 6-10 years | 6 |
|  |  | >10 years | 6 |

|  |  |  |
| --- | --- | --- |
| Gender | Female | 23 |
|  | Male | 17 |
| Ethnicity | White British | 33 |
|  | British Asian | 4 |
|  | Not reported | 3 |

We completed a total of 162 hours non-participation across the six ED sites (see table 3 for details).

**Table 3:** Details of pre-alerts observed.

| ED |  | No. of Pre-alerts observed | Type of pre-alert | No of hours observation |
| --- | --- | --- | --- | --- |
| Major Trauma Centre | A (MTC) | 26 | 21 medical, 5 trauma | 31.5 |
|  | D (MTC) | 28 | 26 medical*, 2 trauma | 35.5 |
|  | E (MTC) | 24 | 19 medical, 5 trauma | 25 |
| Trauma Unit | B (TU) | 6 | 6 medical, 0 trauma | 14 |
|  | C (TU) | 34 | 27 medical, 7 trauma | 28 |
|  | F (TU) | 25 | 25 medical, 0 trauma | 28 |

\* one classed as both medical and trauma

### Findings

We identified significant variation in pre-alert communication processes and areas for improvement that could potentially improve ED processes and enhance care for pre-alerted patients. Importantly, we identified how variation could contribute to frustration and tension between emergency services staff, particularly in the context of high pressure and demand. Communication problems related both to *what* **i**nformation was communicated and *how* it was communicated.

### Efficient exchange of pre-alert information is influenced by individual clinician factors

Ambulance clinicians communicated pre-alert information by calling a dedicated ‘red phone’, generally located within the ED resuscitation area (resus). Different levels of experience amongst ambulance staff making these calls were considered to influence the quality of communication (structure, focus and confidence), as well as how the caller and information were perceived by ED staff answering the phone.

> *A senior crew has a bit more - ‘no, no. I know their numbers are, but they’ve got, I’m telling you, but they’ve got a ruptured triple A until proven otherwise, and so they need to come to you full stop’. I think that takes a level of confidence, seniority, experience to be able to do that. [ED48_Registrar]*

> *I’ve found I get, if I pick up the phone and go ‘hello it’s [name] I’m one of the critical care paramedics’ I get a different response than if I don’t. [AS29_SpecialistParamedic]*

> *I think sounding confident and sounding clear, in a sense, especially if you feel like you’re trying to sell someone to resus, rather than just giving them the information…I think, ultimately, it will affect patient care if you don’t sound confident in your handovers. [AS28_Paramedic]*

Similarly, seniority and experience of ED staff was perceived as important in responding to calls as this informed decisions about patient care for which they were ultimately responsible

> *If I hear the phone I will, and particularly if I’m in resus, I will almost always try and get there…I will look over their shoulder and then often say, ‘right, give me the phone’. I think most people don’t get too annoyed…But at the end of the day, having been in coroner’s court and [I’m the one] has to justify why this has happened on my watch. [ED05_Consultant]*

> *Don’t answer it if you don’t feel confident because you’ll flap and not get the information. [ED11_Nurse]*

> *I think it’s imperative that the right person answers the phone…I will hover over somebody who I don’t think should have answered the phone, but I get that they’ve answered it because nobody else has. [ED17_SeniorNurse]*

### Pre-alerts need to be communicated concisely and quickly

Both ambulance and ED clinicians commented on the value of concise pre-alerts, which enabled them to return to delivering patient care as quickly as possible. Observations and interviews revealed that answering the ‘red phone’ created interruptions to activity within the ED, requiring (often senior) staff to leave tasks (including direct patient care) in order to respond to the call, often having to manage multiple calls within a short space of time.

> *You think sometimes when they’re giving you a long handover you’re thinking, I’m on this phone no one else can get through. [ED4_Registrar]*

> *What you want ideally is you want a nice succinct thing of, this person, they’re coming in with this, the background is this, the observations are this, this is what I’ve done, this is how long I’m going to be. End of. [ED23_Consultant]*

Likewise, ambulance clinicians expressed concerns that waiting for a response to their pre-alert call or a protracted pre-alert conversation impacted on their delivery of patient care. Ambulance clinicians described the significant difficulties of making pre-alert calls when they are alone in the back of an ambulance, travelling at speed and trying to stabilise a sick or deteriorating patient.

> *That’s another thing that annoys me as well. When I’ve got a really poorly patient in front me and they want everything including their inside leg measurement over the phone. And it’s like, I’m the only person in the back of the ambulance, I need to treat my patient, I’ll see you in a minute, bye. I’ll tell you everything else when I get there. [AS36_ClinicalLead]*

> *If you ring for a stroke patient, they sometimes want name, NHS number, and they want a little more of a discussion before you come in. So them calls can last up to five minutes. Which again delays transport. [AS1_NewlyQualifiedParamedic]*

Both Ambulance and ED clinicians expressed frustration at calls being unnecessarily extended, either due to interruptions for clarification or a lack of structure within the call making it difficult to identify key information. This appeared to occur when clinicians lacked a shared understanding of the most pertinent information required and excessive information was being provided or requested.

> *I don’t really give anyone the option to ask many questions on the phone…Questions which I don’t think is relevant…I’m like I can give you this information when I arrive but for now, I’m actively trying to treat this patient. [AS7_Paramedic]*

> *I feel like if you make it go on too long they’re not very interested, but they’re a busy department, as long as you get the pertinent information in there, they can’t ignore it. [AS43_EMT]*

### A shared format for information handover facilitates efficient communication of pre-alert information

Communication difficulties arose due to the lack of shared documentation and differential understanding of how to structure the pre-alert conversation. The various ambulance services and EDs observed used different data collection forms for recording and delivering pre-alert information. Documentation included established information handover tools (e.g. ATMIST, ASHICE, SBAR) and local bespoke information requirements. These documents used different formats and ordering of patient details (e.g. clinical observations) to record and share information. Some EDs used separate forms for trauma or medical pre-alerts, whilst others used one form, with the option to tick whether the call was for trauma or another specified condition (e.g. stroke, sepsis).

The use of different formats to share pre-alert information was identified by both ambulance clinicians and ED staff contributing to interruptions, loss of information, and frustration. Some participants commented on the need for greater consistency in documentation so that both services are ‘and pre-alerts are communicated more efficiently. To facilitate this, one ED had created a form to match that of the local ambulance service. However, ambulance clinicians often transported patients to multiple hospitals and EDs often received patients from more than one ambulance service.

> *When you’re not talking the same language, that becomes a problem. If your receiving hospital have got paperwork like that, in an ATMIST or SBAR format, and you try to give a pre-alert in an opposing format, all of a sudden you get a case of, well, are we talking the same language, along the same lines? Your route from A to B is completely different to their route. [AS30_Paramedic]*

> *The order they read their observations out now don’t match ours again. So, you just, if they’re reading us a string of numbers you have to keep jumping all over the place on the form to put them in on our end…It’s just one of those things where we both keep switching systems and they don’t match up. [ED1_SeniorNurse]*

> *We wrote our forms so that it was the same order as what gets displayed by the [Ambulance service] team. [ED14_Consultant]*

There was also evidence of disparities between individual clinicians in how structured formats were used and differential understanding of which formats should be used for different types of patients. Clinicians described using the format that they felt comfortable with or preferred.

> *Regardless of it being medical or trauma I go for ASHICE, I just feel like it works effectively. [AS12_NewlyQualifiedParamedic]*

> *I use the ATMIST for trauma, and I use ASHICE for medical because SBAR wasn’t a thing when I trained, or if it did exist it didn’t exist pre-hospitally, and I don’t hand over in SBAR because I find it very vague. [AS24_Paramedic]*

> *Nurse in charge was saying that for her she really likes the SBAR format that’s been drilled into her. [SiteE_Obs3a]*

Consistent structured formats were perceived as supporting more focussed and succinct pre-alert call interactions, reducing the amount of superfluous information. The use of a structure also helped the ambulance clinician to reflect on the information and prepare mentally for the call, which is particularly beneficial for less experienced clinicians.

> *I think the acronyms are good, cause it stops you from giving too much and too little information. Sometimes handovers can be a bit convoluted if you’re just reeling it off the top of your head. [AS28_Paramedic]*

> *It doesn’t work when they, as well, when crews don’t follow a pattern. So, you know, having a clear handover process, you know, ATMIST or whatever, you know, is used in other places is really important to structure that. [ED5_Consultant]*

> *When I’m pre-alerting I write ASHICE on a piece of paper, and then I literally write it out like that and then I ring them, if I have time. If I didn’t have time I do it off the top of my head but the one’s I do off the top of my head are never as good as the ones I write out. [AS24_Paramedic]*

### Importance of communication in managing expectations and reducing incivility

Conflicting views on what constitutes a pre-alert and what the pre-alert phone should be used for sometimes led to frustration and tension between clinicians, reflected in less courteous responses and questioning of pre-alert decisions. Examples included when pre-alert calls were made due to protocol requirements rather than clinician concern, or when clinician concerns related to future deterioration whilst waiting in an ambulance queue. These and other calls for advice could be a source of irritation to ED clinicians who often did not consider them appropriate for the pre-alert phone. Ambulance clinicians encountered varying responses from different EDs and clinicians with different thresholds for acceptability of these type of calls.

> *If you’re stood already in A&E and the red phone goes off, as they’re writing stuff down you’ll see the eye rolls. [AS2_Paramedic]*

> *When you stand in A&E and you’re queueing you hear the nurses say sometimes, why are they pre-alerting about this…but I think the other thing is every hospital is different. [AS24_Paramedic]*

> *It depends who you speak to, it depends which team are working in the ED. Literally, which nurse you speak to, which consultant is on that day…and that can tell me how my pre-alert is going to go, sometimes. [AS14_Paramedic]*

ED clinicians valued having the information they required in an order that enabled them to decide quickly on the most appropriate response, which included an early understanding of the main clinical concern. When they did not receive the information concisely and quickly, this often led to probing and asking questions. However, interruptions and questioning were often perceived as disruptive by ambulance clinicians, challenging their clinical competency and delaying transport or patient care.

> *The structure’s there for a reason, and actually probably their questions would be answered if they just listened to what we had to say. But to do that in an assertive but polite way is a bit of a tightrope to walk at times. [AS1_ Emergency medical technician]*

> *They do fire more questions at you at [City1] sometimes. They get me in a bit of a – you know, you can be a bit like “Oh, um” – a bit panicky – because they do fire questions off at you. [AS2_Paramedic]*

> *I genuinely don’t know what it is about [Place 2], there are times where you just don’t feel listened to at all. [AS33_Paramedic]*

Ambulance clinicians sometimes felt they were not being listened to when ED staff interrupted them to ask questions or were simultaneously communicating with a colleague in the background. From an ED perspective, questioning was sometimes necessary to obtain specific information quickly for planning and prioritising resources, in consultation with clinicians in charge.

> *The difficulty is sometimes that when the red phone goes and depending on who it is that’s picking it up, they’re being interrupted aren’t they from their task and it’s probably the 100th time they’ve been interrupted that shift. And depending of the quality of the handover or the information delivered maybe they have received a less than warm reception…it shouldn’t matter, should it. Should be civil to one another and listen to each other. [ED23_Consultant]*

> *We are constantly busy and I think we constantly don’t have enough beds to meet demand, and so what we are just trying to do is we are just trying to get as much information as possible to make an informed decision…So I think we ask more questions because we’re trying to get it right because we know we are tight for space. [ED48_Registrar]*

### Need for improved communication and understanding between ED and ambulance services

Different expectations of the information handover combined with limited understanding of each other’s situation also created interruptions and frustrations. With the exception of calls for advice, most pre-alert calls were for patients who ambulance clinicians felt required a resus bed. However, the ED response regarding where the patient should be taken was principally based on resource availability at the time of the call and access to resus was not always possible. Ambulance clinician accounts of these communications indicated that they could perceive these responses as a ‘rejection’ of their pre-alert and questioning of their clinical judgement, particularly when a reason was not provided.

> *‘They are pretty stressed out in [city3]. So, as soon as that phone goes off, anybody now knows that they’re gonna get it…They don’t like putting anyone in resus in [City3]. I think sometimes they think we are exaggerating a situation. [AS02_Paramedic]*

We observed instances where ED clinicians did explain the decision in terms of capacity and reassured the ambulance clinician that they would review the decision if necessary. For less experienced clinicians this avoids assumptions that their pre-alert was unnecessary and potential impact on future pre-alert decisions. A number of ED clinicians also commented that they would prefer to know about ambulance clinician concerns and be prepared than not.

> *We’re full in resus but we’ll find you some space somewhere. [SiteD_Obs1a]*

> *We are well over capacity but we will try and make space so come into resus bit don’t be surprised if we can’t make space. [SiteE_Obs2b]*

Shared expectations of the communication process were more likely in the case of established relationships between individual ambulance and ED clinicians, where mutual trust existed. Inevitably however, this was less likely for newer staff or when ambulance clinicians were conveying to EDs that they visited less frequently.

> *Having worked for the ambulance service for a while, I know staff at these EDs, and it is significantly easier when you know the person and you have a rapport with them. And I guess they also, my colleagues there, understand that I’m not a panicker or a stresser; I don’t put pre-alerts in unnecessarily. [AS30_Paramedic]*

> *…with that familiarity comes a little bit of respect. [AS42_SpecialistParamedic]*

ED clinicians expressed frustration when they were not informed that patients had deteriorated, or when they perceived that ambulance clinicians had exaggerated the patient’s condition to get seen quickly. For ED clinicians having an update on the patient’s condition meant they could ‘step down’ the pre-alert in the event of a sudden deterioration. Different views on the need to call about patients that had stabilised or deteriorated very close to arrival at the hospital were also a source of tension.

> *The only thing that I would say is quite frustrating, is once a crew has alerted, especially if it’s someone that’s almost peri-arrest, there have been a few occasions where they haven’t called us back to say they’re actually now in cardiac arrest, or this patient has significantly deteriorated. I suppose they’re busy on the back of the truck with the patient, but it’s quite frustrating that they don’t keep us up to date from that phone call. [ED33_SeniorNurse]*

### Training is needed to deliver more consistent communication in pre-alert calls

Despite acknowledgement of the importance of consistent pre-alert practice, this was not a key skill addressed in emergency clinician training. Both ambulance and ED clinicians reported minimal training on pre-alerts, learning how to undertake and receive pre-alerts by watching and listening to colleagues, often receiving guidance from more senior clinicians during the pre-alert.

> *There’s no formal training, it’s all informal training… I think it’s a under recognised skill. So to take a good call… If you’ve not asked the right questions, actually that’s sometimes just as bad as interrupting someone. [ED43_Consultant]*

> *We had an induction, which said we should answer the phone, they showed us the sheet and then said if you have any questions at all there should be a registrar or a consultant nearby. [ED35_Doctor]*

> *You pick up the phone and get passed a piece of paper and off you go. There isn’t really much to do…I haven’t even thought of training somebody to do it. [ED14_Consultant]*

Most ambulance and ED clinicians identified the need for training to improve consistency of communication to ensure that staff in both services have a shared understanding of the purpose and conduct of the pre-alert call. The requests for training reflected a general desire for more standardisation of pre-alert communication both within and across the services, as discussed above.

> *I think there’s a basic kind of bit of training that needs doing around how to have those conversations [pre-alert call]…Which is ‘this is what I want from you from this call, this is the information I’m giving you, this is my plan’. [AS29_SpecialistParamedic]*

> *I think that nationally, across the board, it would be good for alert call training and it to be very standardised. [ED17_SeniorNurse]*

> *Where the variation comes I think is at our end…You get a wide variety of people, doctors, nurses, ODPs, senior, junior, answering the phone, I’m not sure that any of them have actually had any training in how to do that, I certainly haven’t. You know, no ones ever taught us this is what we need to know. [ED38_Registrar]*

### Early communication of key information provides clarity and helps to frame the pre-alert

Observation and interview data demonstrated the importance of basic information being shared at the beginning of the call, including introductions (on both sides), an estimated time of arrival and brief patient details. Ambulance clinicians also valued ED staff answering the phone by stating who they were and the name of the ED, partly to ensure they had the correct ED but also to ‘cover their backs’ in the event that the information did not get passed on. Ambulance and ED staff recounted relatively rare occurrences of ambulance staff unknowingly pre-alerting the wrong hospital, which may have been avoided if the ED was identified at the outset.

> *Often you’ll get an answer of “Hello, A&E” and you’re like, Which A&E is that?… it’s years ago. I pre-alerted the wrong hospital. [AS30_Paramedic]*

> *There is the odd, the rare occasion…You get to a hospital and they’re not expecting you and you say, “Well somebody made a pre alert, I heard them doing it …” And then you find out that they rang a different hospital, so they’ve got an entire team waiting at a hospital that you’re not at. [AS46_SpecialistParamedic]*

> *We’ve had some that are sort of 20 – 25 minute calls… almost a bit too much notice. And then we’ve had some that are like ‘I’m outside the hospital. We’re just turning into the hospital site now’. And it’s like ‘ah right, well we’ve gotta do a move around in here before you can get in!’. So it’s not enough notice. [ED55_SeniorNurse]*

ED clinicians valued being given a ‘headline’ concern or reason for the pre-alert call to frame subsequent information. Understanding this initial concern was considered helpful for ED staff to interpret the information being given and focus further questions. Although useful in structuring and focussing handover of observations, some ambulance clinicians considered that checklists were limiting in their focus on observations and not allowing for concerns relating to clinical gestalt where observations were felt to either over-or under-estimate the seriousness of the patient’s condition (i.e. ‘they just don’t look right’).

> *I sort of say ‘what is worrying you about this patient?’ because then that kind of gets them to really focus down and it’s almost a bit like ‘oh ok what is the bit that I’m most worried about? [ED51_Practitioner]*

> *Say it’s one of the grey ones, where there is maybe a bit of complexity to it or something like that, I tend to just give a bit more of an explanation as to why. Because sometimes just giving a set of observations can sort of give the wrong impression of a patient. [AS18_Paramedic]*

### Available practical resources and technologies impact on pre-alert communication

Clear communication was hindered by a number of practical problems, where technological solutions were not yet available. Ambulance services used different communication methods, with only one of the three services operating recorded radio lines for clinicians to communicate pre-alert information. Provision of work mobile phones and iPads/tablets was variable and ambulance clinicians described having to use their personal mobile phones rather than work phones or recorded lines due to poor signal, particularly in rural areas. Practical difficulties such as loss of signal or poor quality communication lines meant that calls were sometimes cut off before key information was shared.

> *The line is really bad. It’s awful. There’s a big delay on it and it’s often quite quiet and the department’s noisy. So, it’s [a] very challenging conversation when you’re having to ask for information. [ED18_Registrar]*

> *They put this in the ambulance, so the ARP [Ambulance Radio Programme] radios. What I tend to do more often than not is actually just pick up my mobile phone and use my phone…I think there’s better reception on a mobile phone. So, I know we’re always advised to have that call recorded and use the ARP but a lot of the time the call quality isn’t good enough. [AS16_Paramedic]*

Practical changes were suggested, including a second ED phone line to avoid delays when busy and improving the sound or signal quality on phones/radios. An ‘amber’ phone line for pre-alert related advice was considered of value in the current context of ED delays and the risk of deterioration, though ED staff felt that advice should come from within the ambulance service.

> *A second phone would be good. Or an alternative phone number so if the first one was, like I said if you’re on hold, then you have an alternative way to contact them. [AS5_NewlyQualifiedParamedic]*

Technology available for transferring additional information varied across ambulance services and EDs, with clinicians in some services being able to share electronic patient records and ECGs with the ED in advance of arrival. This facilitated access to patient information beyond that communicated in the pre-alert call. The need for greater use of call recording technology was also highlighted, to ensure that all pre-alert call information is available for review later.

> *Fast ECG, the app – which is fantastic. You snap a photo of a myocardial infarction, gives you a code, you send it off and then give them the code on the pre-alert and they can look at it. While you’re talking and giving a history, they can be looking at the ECG. But [Location 2] don’t do that – they want a verbal description of the ECG and what you’ve found to be concerning. […] you’re wasting a lot of time talking to them and trying to describe what you’re seeing. [AS37_Paramedic]*

> *We can see their EPR’s before they arrive and we can see them on the CAD [computer aided dispatch] actually if we need to we can look at most of the stuff. So I can look at patients obs, we can look at their ECG.[…] where it sounds like this is a complex patient, someone that we know a lot, especially in paeds. Or if it’s been a bad line and we’re going to get an idea and then have a look at their notes. [ED40_Registrar]*

## Discussion

We identified sources of variation in pre-alert communication that influenced the efficiency of pre-alert communication, with implications for patient care and the working relationships between emergency services staff. Differences in pre-alert communication formats and clinician expectations undermined the common aspiration for concise pre-alerts and was a source of tension for both ambulance and ED clinicians. Beyond the interpersonal aspects of pre-alert communication, variation in access to communication technologies and ED resource constraints contributed additional practical challenges. Support for a more consistent approach to documentation and what information needs to be communicated was emphasised by ambulance and ED staff, and particularly information that needed to be prioritised in the event of practical communication problems.

Existing literature about communication during the prehospital and ED interface relates primarily to patient handover (24) (25), and supports some of the findings within our study. Other studies have identified variable quality of information at handover (12-14). Bost et al observed that handover practice was learnt ‘on the job’, relying on peer observation, and reported similar issues relating to trust in the information handed over, with good working relationships between ED and ambulance staff leading to improved handover of care.

Fitzpatrick et al similarly identified that ‘interruptions’ were considered the significant barrier to effective prehospital to ED handover and identified the importance of developing a shared mental model through system standardisation(26). Their development of a prehospital/ED shared tool for pre-alert and handover was found to be feasible and identified potential improvements to the recording of important clinical variables, although follow-up was limited. (27)Other studies have similarly advocated for improved consistency in data handover and a need for structured communication procedures and feedback, with uniform patient handover practices both within and across organisations. (28, 29) Dúason et al. identified that a lack of structured communication procedures and feedback as well as ambiguity about patient responsibility in patient handovers from EMTs to ED healthcare professionals may compromise patient safety. Whilst lessons may be learned from patient handover, there are limitations to transferability of findings due to the different purposes of pre-alerts and handover, and the lack of face-to-face communication in pre-alerts.

Clinician experience was identified as important to good quality pre-alert communication within our fieldwork but there appeared to be a lack of training to meet this needEvans et al identified that although paramedics were handing over more information than could be retained, ED staff may be able to improve information recall by improving active listening. This supports our finding that training should be improved for both delivering (ambulance clinicians) and receiving (ED clinicians) pre-alerts. Wilson et al. highlight the potential benefits of feedback for clinical practice, patient outcomes and staff mental health. In common with the findings from this study, they identify lack of feedback as an issue for ambulance clinicians in the UK and internationally, despite an expressed desire for such information.(29)

We identified that limited understanding of joint perspectives could lead to frustration and incivility. Incivility has been shown to cause immediate and short-term affective responses as well as longer-term reduction in mental health (30) Incivility has been shown to pose serious risk to communication and information sharing, leading to erosion of quality and safety of patient care. (31, 32) Cash et al reported that almost half of EMS workers experienced incivility at least once a week, with significant impact on morale and job satisfaction. (33) Credland et al explored paramedic perspectives of incivility and highlighted that incivility from other colleagues or professional groups was most challenging, impacting how they work and subsequent clinical decision-making. (34) This aligns with our finding that ambulance clinicians were conscious of incivility and perceived judgement of the appropriateness of their pre-alert and its potential impact on their future decision-making. Improved communication by ED staff of their pre-alert response (i.e. explaining why patients are not being taken to resus), and active listening during pre-alert conversations may reduce perceptions of incivility and lead to improved ED-ambulance clinician relationships.

Practical issues relating to the quality of pre-alerts were corroborated within a UK national survey of ambulance clinicians, which identified that 46% of ambulance clinicians use their personal mobile due to problems with connectivity and signal. (17) Bost et al similarly identified that telephone communication of critical cases was poor due to poor sound quality. (24) Technological difficulties accentuate the need for a clear, structured pre-alert with a headline concern to mitigate for potential loss of phone signal.

### Limitations

We observed pre-alert practice from the ED perspective but were unable to observe the calls from the ambulance perspective due to the small number of pre-alerts occurring in each shift. We may have been able to identify further potential areas of improvement had we had the opportunity to observe ambulance clinicians undertaking pre-alerts.

Our fieldwork took place mainly within larger EDs (Major Trauma Centres or Trauma Units) rather than minor units where pre-alerts occur less frequently and may be managed differently. Fieldwork also took place within ambulance services where the majority of pre-alerts were undertaken directly between ambulance clinicians and ED staff and findings may be less transferable to ambulance services where pre-alert calls go via a central desk that communicates the information to EDs. Although this research only involved three ambulance services, our findings were reflected in complementary evidence from our linked survey of all ambulance services in England. (17)

### Implications

Our findings suggest that consistency in practice may be improved by greater standardisation of communication tools, training and feedback, and particularly cross-service collaboration to minimise potential sources of tension. The use of a shared format including a headline ‘cause for concern’ may improve the clarity, usefulness and civility of pre-alerts. Basic training on how to undertake pre-alerts for both ED and ambulance clinicians may improve understanding of the importance of pre-alert communication, including the need for ED clinicians to explain their decisions briefly to ambulance clinicians.

Simple pragmatic improvements include the extension of technologies to reduce reliance on communication via phones or radios with poor signals and sound quality. Similarly, identifying the clinician name and ED at the beginning of the call, an ETA and basic summary of clinical concern may mitigate problems caused by practical difficulties such as poor reception or loss of contact during the call.

## Data Availability

The data generated for this study is in the form of confidential transcripts of interviews that are not available for sharing. Participants consented for anonymised quotations to be shared but did not consent to share the full transcripts.

## Acknowledgements

The authors would like to thank all research participants and ED and ambulance service staff who helped to recruit participants. We are also grateful for the input of other members of the study team, our advisory group and our patient and public involvement representatives/group. Thanks to Marc Chattle for clerical support.

